# Los Angeles County SARS-CoV-2 Epidemic: Critical Role of Multi-generational Intra-household Transmission

**DOI:** 10.1101/2020.10.11.20211045

**Authors:** Jeffrey E. Harris

## Abstract

We studied the incidence of confirmed COVID-19 cases diagnosed during February 24, 2020 -January 10, 2021 in approximately 300 communities making up Los Angeles County, the largest county by population in the United States. The surge in case incidence observed from October 19 onward, accounting for two-thirds of all confirmed cases, was concentrated in communities with a high prevalence of multi-generational households, as gauged by data from the American Community Survey. This indicator (abbreviated MULTI) was a more important predictor of the surge in incidence than the prevalence of households with low income or with at least one high-risk worker. Serial mapping of the epidemic revealed radial expansion from an initial focus in relatively affluent communities, followed by concentration in high-MULTI communities. This observation was supported by estimates from a spatial adaptation of the SIR model, which yielded a reproductive number of 2.7 for the initial outbreak during February 24 -March 30. With the subsequent flattening of the epidemic curve after the imposition of emergency stay-at-home orders, the global reproductive number fell to 1.0, but with wide local dispersion ranging from 0.6 in low-MULTI communities up to 1.5 in high-MULTI communities. The July 13 state-ordered reversal of the county’s prior decisions to reopen retail stores, indoor dining, hair salons, gyms and bars had a larger negative impact on social mobility in high-MULTI communities, as gauged by data from SafeGraph on smartphone visits to fast-food restaurants. After falling to a low of 0.6, the reproductive number rebounded to 1.4 during the final surge. By the end of the 46-week observation period, the estimated cumulative incidence of COVID-19, adjusted for underascertainment of both asymptomatic and symptomatic cases, ranged from under 10 percent in low-MULTI communities to over 30 percent in high-MULTI communities.

## 1. Introduction

In this article, we attempt to identify the critical forces driving the massive outbreak of COVID-19 in Los Angeles County, which by January 10, 2021 had registered over 967,000 confirmed cases of the disease (Los Angeles County Department of Public Health 2021).

To that end, we bring together four critical strands of the growing research literature on the worldwide COVID-19 epidemic. First, investigators have attempted to reconstruct the transmission dynamics of local outbreaks by applying theoretical models to data on reported cases (Hao et al. 2020, Fang, Nie, and Penny 2020, Chang et al. 2020). Second, numerous studies have used the techniques of geospatial analysis to evaluate the impacts of public health policies (Franch-Pardo et al. 2020, Dickson et al. 2020, Orea and Alvarez 2020, Zheng et al. 2020). Third, cross-sectional studies have related the age structure and household composition of various countries to COVID-19 incidence and mortality (Esteve et al. 2020, Aparicio Fenoll and Grossbard 2020). And fourth, researchers have increasingly relied on data derived from the movements of smartphones with location-tracking software to study patterns of viral propagation (Dave et al. 2020, Harris 2020a, d).

While a number of studies have assessed the effects of state-of-emergency and stay-at-home orders, as well as restrictions on restaurants, bars and large social gatherings, these efforts have largely relied upon large cross-sections of state and county data (Cronin and Evans 2020, Gupta et al. 2020). Here, by contrast, we rely upon detailed data on the dynamics of SARS-CoV-2 transmission among approximately 300 communities within Los Angeles County from February 24, 2020 through January 10, 2021. Focusing sharply on Los Angeles County – far and away the largest by population in the United States – we follow in the line of other recent studies attempting to relate transmission patterns to the fine microdetails of individual communities (Vijayan et al. 2020, Horn et al. 2020).

We develop a spatial extension of the conventional SIR epidemic model to study the radial spread of infection among these contiguous communities during the early phases of the epidemic. We merge our geospatial data with census-derived, community-specific data on the characteristics of households, in particular a measure of the prevalence of households at risk for inter-generational transmission. We study how wide local variations in the prevalence of this risk factor result in marked heterogeneity in the reproductive number during the course of the Los Angeles county epidemic.

### 1.1. The Four Phases

Before delving into the fine details of our data, methods and results, we paint a broad-brush picture of the epidemic under study.

Figure 1 plots the weekly incidence of confirmed COVID-19 cases per 100,000 population in Los Angeles County over a 46-week period, from the week starting Monday, February 24, 2020 (which we designate as week 0) to the week starting Monday, January 4, 2021 (designated week 45). The data points for the figure were derived principally from the web-based dashboard of the Los Angeles County Department of Public Health (DPH), supplemented by the dashboards of the cities of Long Beach and Pasadena, which are situated within Los Angeles County but run their own health departments (Los Angeles County Department of Public Health 2021, Long Beach Department of Health and Human Services 2021, Pasadena Health Department 2021). We characterize these as *confirmed* cases, as they reflect only those infected individuals who were tested and reported to the DPH or to the two other municipal health departments.

**Figure 1.**
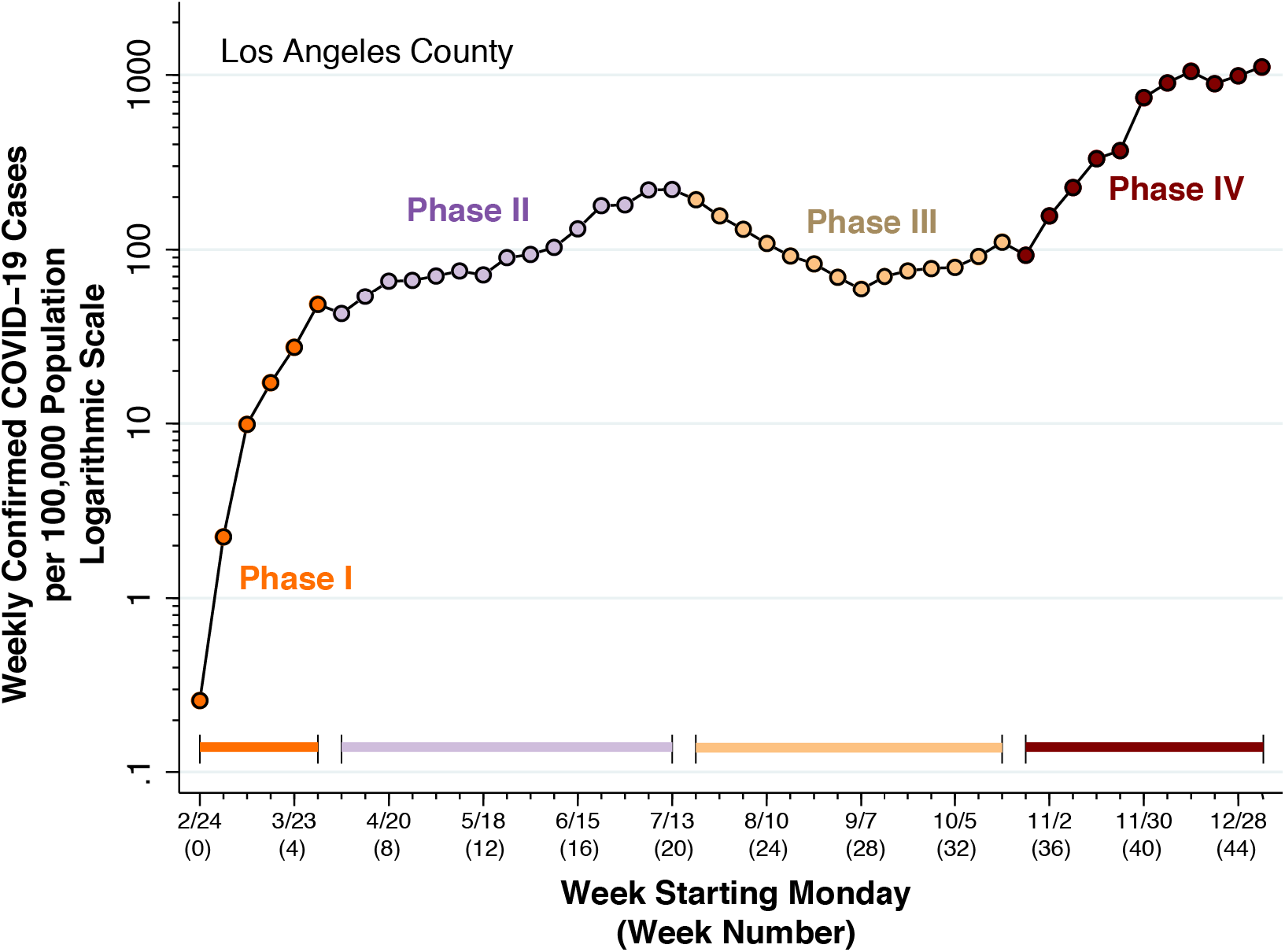
Weekly Confirmed COVID-19 Cases per 100,000 Population in Los Angeles County

We have divided the observation period into four successive phases. *Phase I* (spanning weeks 0–5) saw an initial rapid increase in confirmed case incidence. By the start of *Phase II* (spanning weeks 6–20) the epidemic curve was already flattening, as the emergency lockdowns declared in early March by the Los Angeles mayor, the county supervisor and the California governor had begun to bite (Garcetti 2020a, Barger 2020, Newsom 2020). Following the reopening of retail stores, indoor dining, hair salons, gyms and bars in May and June (Parvini 2020, Shalby 2020a, Money 2020), confirmed case incidence rose to a temporary peak of 221 per 100,000 by the week of July 13 (week 20) at the end of Phase II.

Reacting to the surge in cases, the state public health officer ordered the reversal of most of the county’s prior reopening orders (Gutierrez 2020, Angell 2020). The resulting decline in confirmed case incidence, seen during the initial part of *Phase III* (spanning weeks 21–34) was short-lived, as the county reopened hair salons, nail salons, breweries and shopping malls in September and October (Cosgrove 2020, Shalby 2020b, Shalby and Cosgrove 2020). As Phase III came to a close in week 20, the state issued new guidelines permitting gatherings of up to three households (Times Staff 2020).

The extraordinary, ten-fold surge in confirmed case incidence that followed during *Phase I*V (spanning weeks 35–45) elevated Los Angeles County to the title of the new COVID-19 epicenter in the United States. Emergency stay-at-home orders issued by the Los Angeles mayor and the California regional health officer during week 40 (Garcetti 2020b, Pan 2020) had little or no short-term detectable effect on indicators of social mobility (Harris 2020f). Confirmed cases during the eleven weeks of Phase IV accounted for more than two-thirds of all confirmed cases recorded during the entire 46-week interval covered by the figure.

## 2. Data and Methods

### 2.1. Countywide Statistical Areas

We have already noted our data sources for confirmed COVID-19 cases in Los Angeles County and the cities of Long Beach and Pasadena. While Figure 1 shows incidence trends at the global, countywide level, our analysis was focused principally on a detailed study of confirmed case incidence among the more than 300 communities within the county.^*^ For this purpose, we relied upon the DPH’s geographic breakdown based upon *countywide statistical areas* (CSAs), a mixed classification of independent cities such as the City of Beverly Hills, neighborhoods within the city of Los Angeles such as Hollywood, and unincorporated places such as Hacienda Heights (City of Los Angeles 2020). In this scheme, both Long Beach and Pasadena have their own CSAs.

### 2.2. American Community Survey Data

We relied on the 2015–2019 five-year public use microsample from the U.S. Census Bureau’s American Community Survey (ACS) (U.S Census Bureau 2021). The nationwide database covered 788,475 households and group living arrangements with a total of 1,887,461 persons. Out of the entire database, 203,545 households and group living arrangements with 510,501 persons were identified as residing in one of 69 public use microdata areas (PUMAs) within Los Angeles County (U.S Census Bureau 2020b).^†^

We used the person records of the public use microsample to identify households with at least 4 persons, of whom at least one person was 18–34 years of age and at least one other person was at least 45 years of age. We describe these households here as *at risk* for multi-generational transmission (abbreviated *MULTI* in the results below). Among all such at-risk families in the Los Angeles County extract of the ACS, 43 percent had 4 persons, 27 percent had 5 persons, 14 percent had 6 persons, and 15 percent had 7 or more persons. Among the 69 public use microdata areas (PUMAs), the median proportion of at-risk households was 14.6 percent.

Using the internal household sampling weights provided by the ACS, we then computed the proportion of at-risk households in each PUMA. Applying a Census Bureau crosswalk between PUMAs and census tracts (U.S Census Bureau 2020a) as well as a DPH-provided crosswalk between census tracts and CSAs, we determined the corresponding proportions of at-risk households in each CSA. Among 300 CSAs, the median proportion of at-risk households was 13.8 percent.

We similarly used the ACS five-year public use microsample to determine three other CSA-based indicators: the proportion of households receiving food stamps under the Supplemental Nutrition Assistance Program (*SNAP*, median 7.3% among 300 CSAs); the proportion of households with total income below $22,000 annually, the amount that one full-time worker would earn at California’s minimum wage of $11 per hour (*INC22*, median 15.5%); and the proportion of households with at least one person engaged in a low-wage occupation that cannot be performed remotely (*OCCUP*, median 14.1%).^‡^

### 2.3. SafeGraph Data

We relied upon the Patterns database issued by SafeGraph (SafeGraph Inc. 2020), which describes the movements of smartphones equipped with location-tracking software to numerous *points of interest* throughout the United States. We previously relied upon this data source in a comparative study of the COVID-19 epidemics in Milwaukee and Dane Counties in Wisconsin (Harris 2020a) and a geospatial analysis of the September 2020 COVID-19 outbreak on the campus of the University of Wisconsin-Madison (Harris 2020e).

Here, we focused on fast-food restaurants as points of interest. We used the Patterns *location_name* variable to identify all entities whose names included at least one of these key words: burger, pizza, pizzeria, taco, taqueria, quesadilla, burrito, chipotle, tortilla, sushi, sashimi, ramen, udon, wok, and noodle. We then used the Patterns *brands* variable to identify other fast-food chains that were prevalent in Los Angeles County.^§^

Many of these restaurants, particularly the chains, had multiple locations. Each distinct location was identified by a point-of-interest census block group (*poi_cbg*). For each distinct location, we used the variable *visitor_home_cbgs* to identify the home census block groups of all visitors during each weekly reporting period, where a device’s *home* is the location where it is regularly located overnight. For each week from the week starting February 10, 2020 (week 0) through the week starting January 11, 2021 (week 45), we then accumulated the respective numbers of restaurant visits originating from each home CBG. Once again taking advantage of the census tract crosswalk provided by the Los Angeles County DPH, we converted these counts into a longitudinal time series of restaurant visits originating from each CSA.

### 2.3. Spatial SIR Model

We devised a spatial adaptation of a discrete-time SIR (susceptible-infective-resistant) model similar to the model we employed in a study of COVID-19 transmission between younger and older persons in Florida’s most populous counties (Harris 2020b).

To facilitate the exposition, we first review a deterministic SIR model without a spatial component. Let *S*_*it*_ denote the proportion of susceptible individuals in geographic unit *i* at discrete time *t*. In our empirical application, geographic units will refer to CSAs, and each time period *t* will refer to one week, where *t* = 0, 1, …, 45. Let *I*_*it*_ denote the corresponding proportion of infective individuals, and *R*_*it*_ the corresponding proportion of resistant individuals. For each CSA *i*, the equation of motion of *S*_*it*_ is given by

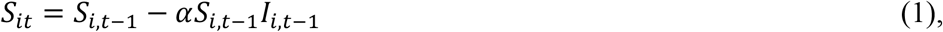

where *α* reflects the rate at which susceptible and infective individuals interact, as well as the likelihood that an interaction will result in transmission. We take *α* to be an unknown parameter to be estimated from our data.

The corresponding equation of motion of *I*_*it*_ is given by

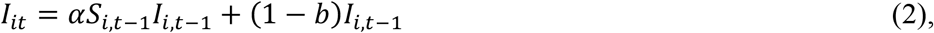

where 1 > *b* > 0 is the rate at which infectives become resistant, either through recovery or death, and (1 − *b*) is the corresponding depreciation factor. Rather than estimating *b* from our data, we rely on external sources. With a mean duration of infectivity of 5.5 days (Griffin et al. 2020), we assume a weekly depreciation factor of 1 − *b* = *exp*(−7/5.5) = 0.28, that is, *b* = 72 percent of current infectives become resistant each week. In sensitivity analyses, we tested the effect of increasing the mean duration of infectivity to 6.5 days, so that 1 − *b* = *exp*(−7/6.5) = 0.34, that is, *b* = 66 percent of current infectives become resistant each week. We further assume that the population of each CSA is closed, so that

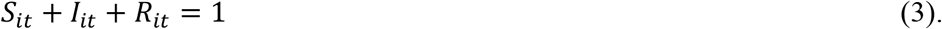

Finally, for each CSA *i*, we assume the initial condition *S*_*i*0_ = 1 − *I*_*i*0_, where *I*_*i*0_ > 0 denotes the proportion of infectives during the initial week *t* = 0, and where *R*_*i*0_ = 0.

We now add a stochastic component to our deterministic model of equations (1) through (3). For notational compactness, we write *y*_*it*_ = *S*_*i,t*-1_ – *S*_*it*_ as the *incidence* of COVID-19 cases in CSA *i* during week *t*. We also write *X*_*it*_ = *S*_*it*_*I*_*it*_. Equation (1) can then be written as

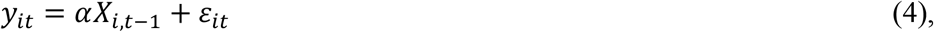

where the error terms *ε*_*it*_ are assumed to be independently and identically distributed. We designate this specification as *Model 0*. This model excludes a constant term, which is ordinarily included in linear models, because all new infections are assumed to arise from contact with other infective persons.

So long as we take the parameter *b* as known, we can estimate the unknown parameter *α* in equation (4) from the available data {*y*_*it*_} on COVID-19 incidence in each CSA *i* and week *t*. That information is sufficient to generate the entire series of *X*_*it*_. To that end, we start with *I*_*i*0_ = *y*_*i*0_ for all *i*, so that *S*_*i*0_ = 1 − *I*_*i*0_ = 1 − *y*_*i*0_. For all subsequent weeks *t* > 1, we compute *S*_*it*_ = *S*_*i,t*-1_ – *y*_*it*_, and then generate the values of *I*_*it*_ from equation (2). Once we have computed *S*_*it*_ and *I*_*it*_, we have *X*_*it*_ as well. We note that the lagged values *X*_*i,t*-1_ in equation (4) are not constructed from the contemporaneous incidence *y*_*it*_ on the left-hand side.

We now incorporate a spatial component into our non-spatial Model 0. We write

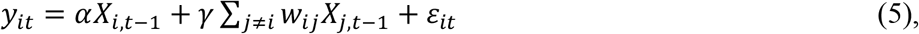

where {*X*_*j*,t-1_: *j* ≠ *i*} refers to all other CSAs, and where both *α* and *γ* are unknown parameters. Here, {*w*_*i,j*_} are elements of a known symmetric matrix *W* with zero diagonal elements, where each off-diagonal element represents the influence of geographic unit *j* ≠ *i* on the rate of new infections in unit *i*. The parameter *γ* thus captures the influence of nearby CSAs. In our empirical application, we set the off-diagonal element *w*_*ij*_ = 1 if the distance *d*_*ij*_ between the centroids of CSA *i* and CSA *j* was no greater than the radius *r*, and *w*_*i,j*_ = 0 otherwise, where the distances *d*_*ij*_ were calculated from the Haversine formula (Hedges 2002). In our base case, we specified a radius *r* of 1 km, but we also studied the effect of increasing *r* to 1.5 km. We designate the specification in equation (5) as *Model 1*.

Once again, this model excludes a constant term because all new infections are assumed to arise from contact with other infective persons located within the same or adjacent geographic units. As above, we can estimate the unknown parameters *α* and *γ* from the available incidence data {*y*_*it*_} so long as we take the depreciation rate *b* and and the matrix *W* as known. Similarly, the lagged values *X*_*i,t*-1_ and {*X*_*j*,t-1_: *j* ≠ *i*} in equation (5) are not constructed from the contemporaneous incidence *y*_*it*_ on the left-hand side.

We consider a further extension of Model 1 that permits covariates. In keeping with the strong assumption that all new infections arise from contact with other infectives, this model takes the form

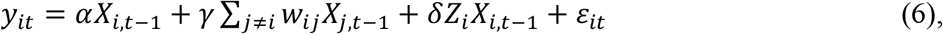

where *Z*_*i*_ represents a time-independent exogenous characteristic of CSA *i*, and *δ* is an additional unknown parameter capturing the multiplicative effect of this covariate on the within-CSA transmission rate. We designate this specification as *Model 2*. In what follows, we estimate Model 2 where *Z*_*i*_ represents the prevalence of at-risk multi-generational households (*MULTI*) in CSA *i*.

Finally, we recognize that the parameters of our models are unlikely to remain constant during the 46-week time period under study. Accordingly, we estimate the parameters *α, γ*, and *δ* separately for each of the four phases described in section 1.1 above.

### 2.4. Calculating the Reproductive Number

In our non-spatial Model 0, our estimate of the contemporaneous reproductive number in CSA *i* at week *t* would be ℛ_*it*_ = *αS*_*it*_/*b*. When ℛ_*it*_ = 1, the equation of motion of the proportion of infective persons gives *I*_*it*_ = *I*_*i,t*-1_. When ℛ_*it*_ > 1, the proportion of infectives is increasing, and when ℛ_*it*_ < 1, the proportion is decreasing. In spatial Model 1, our estimate becomes ℛ_*it*_ = (*α* + *γ*)*S*_*it*_/*b*, where the term (*α* + *γ*) represents the sum of the within-CSA effect and the effect of nearby CSAs. By extension, in spatial Model 2, we have ℛ_*it*_ = (*α* + *γ* + *δZ*_*i*_)*S*_*it*_/*b*. In our empirical analysis below, we estimate the contemporaneous reproductive numbers for the entire county at the starting week for each of the four phases, that is, at *t* = 0, 6, 21, and 35, respectively. To that end, we replace *Z*_*i*_ with the population-weighted mean of *MULTI* for the county and *S*_*it*_ with the population-weighted proportion of survivors at week *t*.

### 2.5. Accounting for Underascertainment of Cases

It is widely recognized that confirmed COVID-19 cases undercount total incident infections. Asymptomatic cases appear to constitute at least 40–45 percent of all infections (Oran and Topol 2020) and appear to play a dominant role in disease transmission (Moghadas et al. 2020). Still other symptomatic individuals may not have sought testing, especially in the early days of the epidemic when testing criteria were restricted (Centers for Disease Control and Prevention 2020, Reese et al. 2020).

A straightforward way to account for such underascertainment is to proportionately inflate confirmed case counts. If *n*_*it*_ is the observed number of *confirmed* cases and 1 > *f* > 0 is the proportion of all infections that go undetected, then the actual number of incident infections would be *y*_*it*_ = *n*_*it*_/(1 − *f*). In our tests of the three spatial models, we applied this inflation factor to confirmed cases, based on the alternative values *f* = 0.4 and *f* = 0.5.

During the earliest weeks of the epidemic, when very few cases have accumulated, nearly everyone remains susceptible, so that *S*_*it*_ ≈ 1 and *X*_*it*_ ≈ *I*_*it*_. In that case, the deterministic version of our model without spatial effects in equation collapses to *y*_*it*_ ≈ *αI*_*i,t*-1_. Inflating confirmed cases by a factor 1/(1 − *f*) would have a negligible effect on our estimates of the transmission parameter *α* and the radial expansion parameter *γ*. As the epidemic progresses, however, the correction for underascertainment will magnify the decline in *S*_*it*_ and thus increase our estimates of *α* and *γ*.

### 2.6. Summary of Critical Parameters

Table 1 summarizes the critical parameters of our spatial epidemic model. In the table, *b* is the proportion of infectives who become resistant each week, as shown in equation (2). The base-case and alternative values were based on assumed mean durations of infectivity of 5.5 and 6.5 days, respectively. The parameter *r* is the radius of influence underlying the definition of the elements {*w*_*i,j*_} in equation (5). In the base case, we take *w*_*i,j*_ = 1, when the distance *d*_*ij*_ between the centroids of CSAs *i* and *j* is no more than 1 km. In the alternate case, we increased the radius of influence to 1.5 km. The parameter *f* is the assumed proportion of all infections that go undetected, which we assumed to be 0.4 or, alternatively, 0.5.

**Table 1.** Spatial SIR Model Parameters

| *Parameters Assumed Known* |  |  | *Parameters to be Estimated* |  |
| --- | --- | --- | --- | --- |
| Parameter | Base | Alternative | Parameter | Models |
| $b$ | 0.72 | 0.66 | $\alpha$ | 0, 1, 2 |
| $r$ | 1.0 | 1.5 | $\gamma$ | 1, 2 |
| $f$ | 0.4 | 0.5 | $\delta$ | 2 |

The unknown parameter *α* in equation (1), which reflects the rate at which susceptible and infective individuals interact, appears in all three models. The unknown parameter *γ* in equation (5), which captures the influence of neighboring communities, appears in Models 1 and Finally, the unknown parameter *δ* in equation (6), which captures the effect of the exogenous covariate *MULTI*, appears in Model 2.

## 3. Results

### 3.1. COVID-19 Incidence in Phase IV versus Prevalence of Multi-Generational Households

We focus initially here on Phase IV because of its quantitative importance. We offer some descriptive tests of the potential role of multi-generation transmission during this critical phase, based upon cross-sectional multivariate regression. In the following section, we proceed to our spatial epidemic model, starting with Phase I.

Figure 2 below displays two color-coded maps of the CSAs of Los Angeles County. The left-hand map shows the geographic distribution of all confirmed COVID-19 cases combined during the 11 weeks of Phase IV, expressed as a percentage of the population of each CSA. The right-hand map shows the corresponding distribution of households at risk for multi-generational transmission, expressed as a percentage of all households in each CSA. In both maps, the color gradient has 7 increments, each corresponding to one septile (or 14.2 percent) of all CSAs.

**Figure 2.**
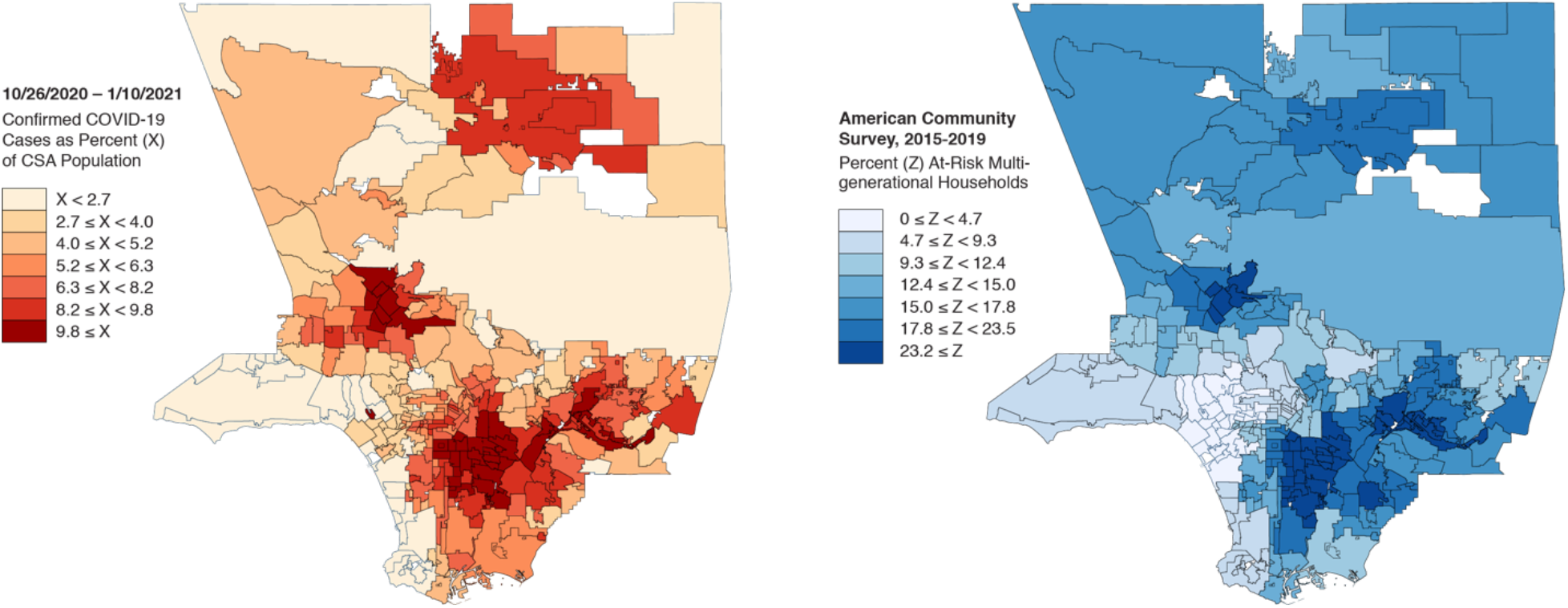
Two Maps of Community Service Areas in Los Angeles County. Left: Incidence of Confirmed COVID-19 Cases Diagnosed during Phase IV. Right: Prevalence of At-Risk Multigenerational Households

Figure 2 shows a striking concordance between the two geographic distributions. Both figures display marked concentrations in four regions: the Antelope Valley–Palmdale–Lancaster region to the north; San Fernando Valley–Pacolma region to the west; the San Gabriel Eastern Valley–El Monte–West Covina–Pomona region to the east; and the Vernon–Boyle Heights–East Los Angeles–Downey–Inglewood region in the center and to the south. On the left, in particular, the two darkest shaded areas (with a cumulative incidence exceeding 8.2 percent) comprised half of all COVID-19 cases during Phase IV but only one-third of the county population.

For the 204 CSAs with a population ≥ 10,000, Figure 3 graphs confirmed COVID-19 incidence during Phase IV against the proportion of households at risk for multi-generational transmission (*MULTI*). Data points are proportional in size to CSA population. The population-weighted least squares fit had a slope of 0.300 (95% confidence interval 0.267–0.334).

**Figure 3.**
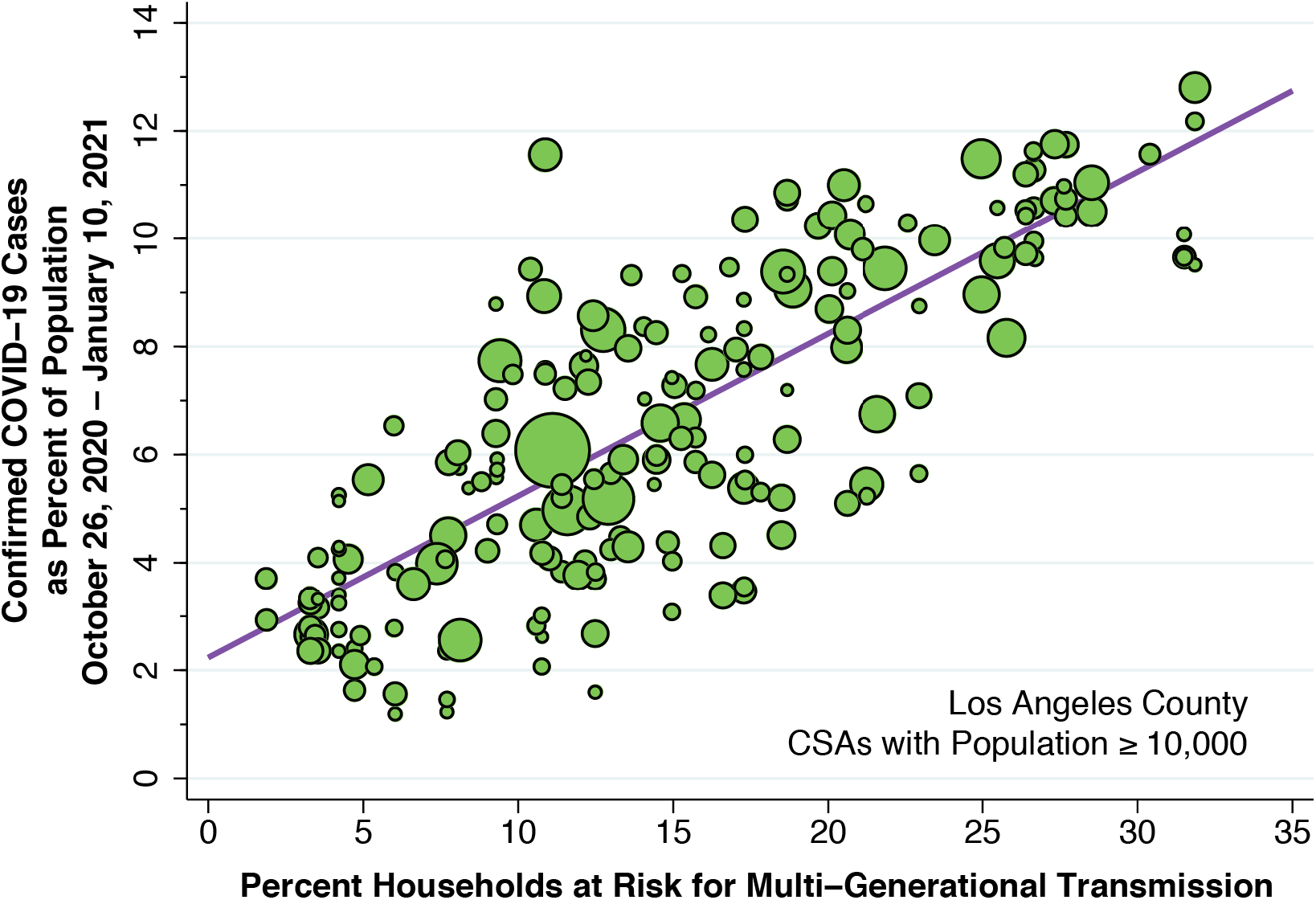
Bivariate Plot of Confirmed COVID-19 Incidence During Phase IV versus the Prevalence of Multi-Generational Households (MULTI)

Table 2 below addresses whether the bivariate relationship between case incidence and the prevalence of at-risk multi-generational households observed in Figure 3 may be attributable instead to other indicators of poverty. The table shows results of population-weighted cross-sectional regressions in two data sets. The first data set, labeled *All CSAs*, covers all 296 CSAs for which we were able to construct estimates for each of the independent variables from the Census Bureau’s American Community Survey 5-Year (2015–2019) database. The second data set, labeled *CSAs* ≥ *10,000*, covers only those countywide statistical areas with at least 10,000 population, as shown in Figure 3 above.

**Table 2.** Cross-Section Regression Analysis of the Percentage of the Population Diagnosed with COVID-19 During Phase IV

| *Regressors* | *All CSAs* | *CSAs $\geq 10,000$* | *All CSAs* | *CSAs $\geq 10,000$* |
| --- | --- | --- | --- | --- |
| *MULTI* | **0.242**<br>(0.022) | **0.246**<br>(0.026) | **0.213**<br>(0.025) | **0.217**<br>(0.029) |
| *INC22* | **0.110**<br>(0.030) | **0.114**<br>(0.035) | **0.106**<br>(0.023) | **0.108**<br>(0.027) |
| *SNAP* | 0.079<br>(0.040) | 0.073<br>(0.046) |  |  |
| *OCCUP* |  |  | **0.090**<br>(0.030) | 0.087<br>(0.034) |
| *Constant Term* | 0.408<br>(0.458) | 0.371<br>(0.537) | 0.265<br>(0.356) | 0.261<br>(0.416) |
| *R<sup>2</sup> Statistic* | 0.705 | 0.720 | 0.710 | 0.722 |
| *Sample Size* | 296 | 204 | 296 | 204 |

Standard errors are shown in parentheses below each parameter estimate. Estimates in boldface were significant at the level $p < 0.01$ , while the estimates in italics were significant at $p < 0.05$ .

Table 2 demonstrates that, while the estimated coefficient of *MULTI* was reduced in comparison with the bivariate model of Figure 3, the prevalence of multi-generational households remained the dominant factor in determining confirmed COVID-19 cases during the Phase IV surge. Inclusion of all four regressors (not shown) resulted in an insignificant relationship for *SNAP* (p = 0.757) and a marginally significant relation for *OCCUP* (p = 0.049). Having offered evidence that *MULTI* dominates over other indicators of poverty, we focus on this variable as the critical covariate in the estimates of the spatial epidemic model below.

### 3.2. Rapid Radial Expansion

The series of six maps in Figure 4 below show the evolution of the cumulative case incidence at the end of weeks 2 through 7, respectively. The lighter shaded CSAs correspond to a cumulative incidence between 120 and 360 per 100,000, while the darker shaded CSAs correspond to a cumulative incidence of at least 360 per 100,000.

**Figure 4.**
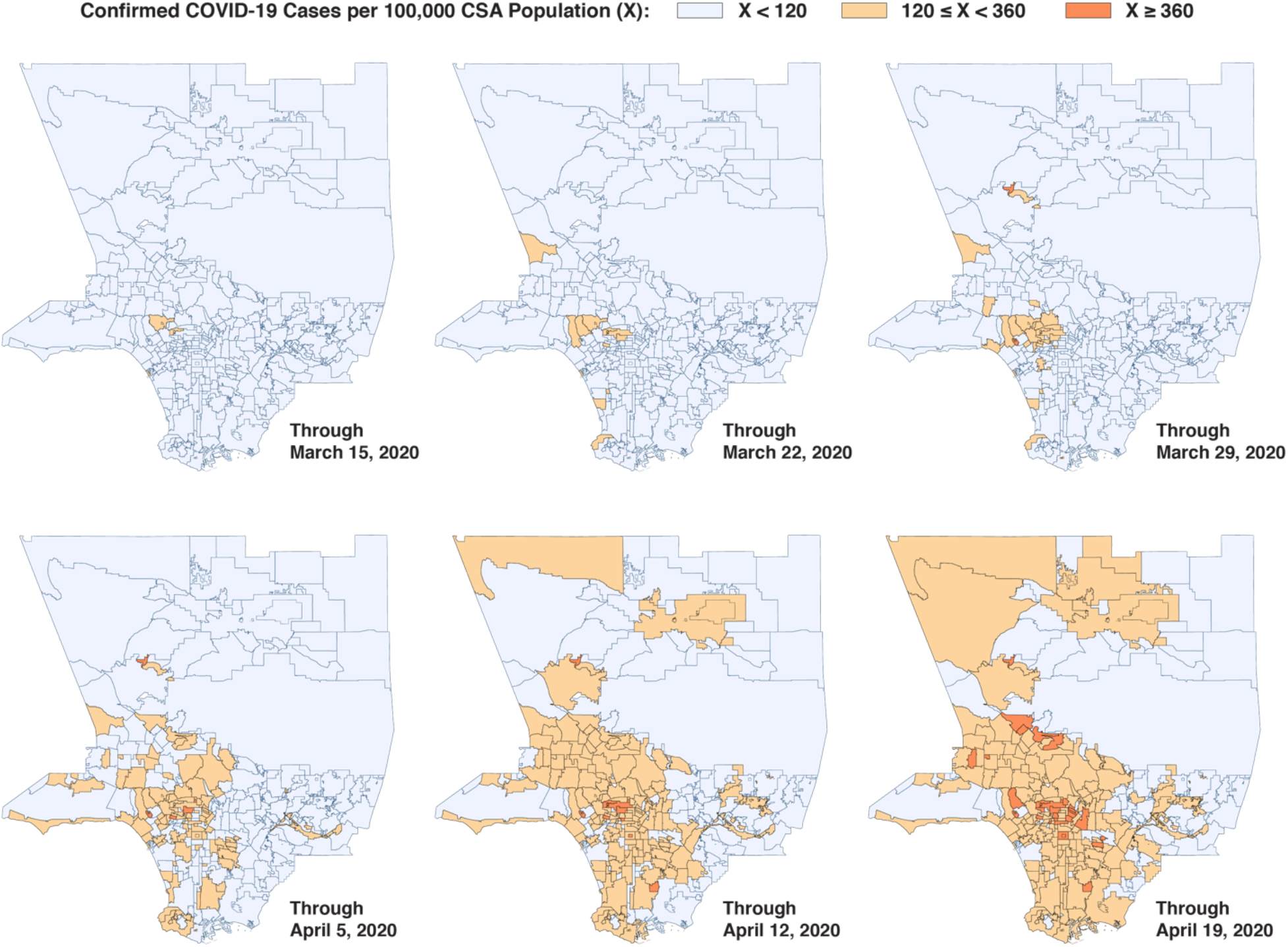
Cumulative COVID-19 Incidence in Los Angeles County at the End of Weeks 3–8

By the end of week 2 (running from March 9–15), a focus of infection exceeding the 120-per-100,000 threshold can be seen in the Beverly Crest community of Los Angeles and the city of West Hollywood. By the end of week 3 (March 16–22), this focus had expanded to include the Brentwood and Belair communities to the west, the Melrose and Hancock Park neighborhoods to the east, and the Crestview community to the south. By the end of week 4 (March 23–29), the focus had further expanded to comprise a cluster of 18 communities, extending to Pacific Palisades to the west, forming a hotspot with cumulative incidence over 320 per 100,000 in the neighborhood of West LA surrounding the Veterans Affairs Medical Center. By the end of week 5 (the conclusion of Phase I), this enlarging cluster added three more communities of high concentration, including Crestview, Hancock Park, and Little Armenia. By weeks 6 and 7 (now in Phase II), the initial focus is no longer distinguishable, and the initial areas of higher concentration had migrated to the south and east.

Based upon Figure 4 alone, the dominant mechanism underlying the continued rapid rise in confirmed case incidence during Phase I appears to be the local radial expansion around a single focus. To be sure, several other isolated areas with a cumulative incidence over the 120-per-100,000 threshold can be seen in such relatively affluent communities as Marina Peninsula and the cities of Manhattan Beach and Palos Verdes Estates to the south. While this observation points to multiple importations by individuals with resources to travel, these parallel importations do not appear to have been controlling.

The map-based observations are supported by the results of Spatial Models 1 and 2 covering Phase I, shown in Table 3 below. We focus first on the base case on the left, where the radius of influence *r* was set equal to 1 km. The significant coefficient for the regressor *W*, which captures the spatial component, supports the radial expansion interpretation. Without any spatial component, the estimated reproductive number came to ℛ = 1.72, but with the inclusion of a spatial component, the estimate increased to ℛ = 2.25.

**Table 3.** Spatial Model Estimates for Phase I (February 24 – April 5)

| *Regressor*<br>*(Parameter)* | Base Case $r = 1.0$ km | | | Alternative $r = 1.5$ km | | |
| --- | --- | --- | --- | --- | --- | --- |
|  | *Model 0* | *Model 1* | *Model 2* | *Model 0* | *Model 1* | *Model 2* |
| $X(\alpha)$ | **1.235**<br>(0.090) | **1.165**<br>(0.092) | **0.775**<br>(0.149) | **1.235**<br>(0.090) | **1.089**<br>(0.094) | **0.669**<br>(0.151) |
| $W(\gamma)$ | | **0.458**<br>(0.126) | **0.467**<br>(0.126) | | **0.289**<br>(0.057) | **0.301**<br>(0.057) |
| $MULTI(\delta)$ | | | **0.047**<br>(0.014) | | | **0.050**<br>(0.014) |
| $\mathcal{R}$ | 1.72 | 2.25 | 2.71 | 1.72 | 1.91 | 2.39 |

Standard errors are shown in parentheses below each parameter estimate. Estimates in boldface were significant at the level $p < 0.01$ , while the estimates in italics were significant at $p < 0.05$ . Sample size = 1,480 in all models. The reproductive number $\mathcal{R}$ was computed at the population mean value of 14.94 for $MULTI$ .

In Spatial Model 2, moreover, the multiplicative effect of *MULTI* was significant. To appreciate the estimate of *δ* = 0.047, consider the effect of a 12-percentage-point increase in the prevalence of at-risk multi-generational households, equivalent to the difference between the population-weighted mean values of *MULTI* in the top and bottom half of the distribution. Since the proportion *S* of susceptible individuals during this early phase of the epidemic is close to 1, the reproductive number would increase by (0.047 × 12)/*b* = 0.78. Turning to the alternative case on the right where the radius of influence *r* was increased to 1.5 km, we see that the estimated parameter *γ* was significantly decreased. This finding supports the conclusion that the influence of nearby communities on the radial propagation of the virus was highly local.

### 3.3. Phase II: Epidemic on the Knife Edge

Table 4, 5 and 6, respectively, display the corresponding parameter estimates for Phases II, III and IV. As in Table 3, each of the tables has two sections. The section on the left shows the base case, while the section on the right shows the effect of varying one of the parameters *b, r*, or *f*.

**Table 4.** Spatial Model Estimates for Phase II (April 6 – July 19)

| Regressor<br>(Parameter) | Base Case $b = 0.72$ | | | Alternative $b = 0.66$ | | |
| --- | --- | --- | --- | --- | --- | --- |
|  | Model 0 | Model 1 | Model 2 | Model 0 | Model 1 | Model 2 |
| $X(\alpha)$ | **0.604**<br>(0.011) | **0.594**<br>(0.011) | **0.203**<br>(0.030) | **0.573**<br>(0.011) | **0.563**<br>(0.011) | **0.199**<br>(0.028) |
| $W(\gamma)$ | | **0.136**<br>(0.026) | **0.197**<br>(0.026) | | **0.123**<br>(0.024) | **0.181**<br>(0.024) |
| *MULTI* ($\delta$) | | | **0.022**<br>(0.002) | | | **0.020**<br>(0.001) |
| $\mathcal{R}$ | 0.84 | 1.01 | 1.01 | 0.87 | 1.04 | 1.04 |

Standard errors are shown in parentheses below each parameter estimate. Estimates in boldface were significant at the level $p < 0.01$ , while the estimates in italics were significant at $p < 0.05$ . Sample size = 4,440 in all models. The reproductive number $\mathcal{R}$ was computed at the population mean value of 14.94 for *MULTI*.

**Table 5.** Spatial Model Estimates for Phase IV (October 19 – January 10)

| Regressor<br>(Parameter) | Base Case $f = 0.4$ | | | Alternative $f = 0.5$ | | |
| --- | --- | --- | --- | --- | --- | --- |
|  | Model 0 | Model 1 | Model 2 | Model 0 | Model 1 | Model 2 |
| $X(\alpha)$ | **0.263**<br>(0.017) | **0.256**<br>(0.017) | **0.710**<br>(0.063) | **0.292**<br>(0.018) | **0.284**<br>(0.018) | **0.707**<br>(0.064) |
| $W(\gamma)$ | | **0.195**<br>(0.066) | 0.059<br>(0.068) | | **0.181**<br>(0.067) | 0.056<br>(0.068) |
| *MULTI* ($\delta$) | | | **-0.020**<br>(0.003) | | | **-0.019**<br>(0.003) |
| $\mathcal{R}$ | 0.36 | 0.61 | 0.63 | 0.39 | 0.62 | 0.64 |

Standard errors are shown in parentheses below each parameter estimate. Estimates in boldface were significant at the level $p < 0.01$ , while the estimates in italics were significant at $p < 0.05$ . Sample size = 4,144 in all models. The reproductive number $\mathcal{R}$ was computed at the population mean value of 14.94 for *MULTI*.

Table 4 shows the parameter estimates for Phase II. The estimated value of the transmission parameter *α*_II_ in the base case is significantly lower than the corresponding value *α*_1_ obtained in the base case in Phase I, as shown in Table 3. (We use subscripts here to distinguish between the estimates for different phases.) This finding is consistent with role of publicly imposed and voluntary lockdowns in reducing transmission during this phase of the epidemic.

While the estimate for *γ*_II_ is still significantly different from zero, the ratio *γ*_II_ / *α*_*II*_ = 0.23 is smaller than the corresponding ratio *γ*_*I*_ / *α*_*I*_ = 0.39 in Phase I. That is, radial expansion continued during Phase II, but had a small quantitative contribution than during Phase I. On the other hand, the ratio *δ*_*II*_/(*α*_*II*_ + *γ*_*II*_) = 0.055 was larger than the corresponding ratio *δ*_*I*_/(*α*_I_ + *γ*_*I*_) = 0.037. That is, transmission via multi-generational households had a larger contribution during Phase II. These conclusions are also borne out in the right-hand panel, based upon an assumed increase in the average duration of infectivity from 5.5 to 6.5 days.

During Phase II, the results for spatial Models 1 and 2 in the base case show the average reproductive number ℛ for the entire county hovering around the knife-edge value of 1. With an estimate of *δ* on the order of 0.02 in Model 2, even a 5-percentage-point increment or decrement in the variable *MULTI* would flip the curve of infectives from increasing (with ℛ = 1.1) to decreasing (with ℛ = 0.9). The resulting heterogeneity of the reproductive number is borne out in Figure 5, which shows the distribution of ℛ_*it*_ at week *t* = 6 at the start of Phase II.

**Figure 5.**
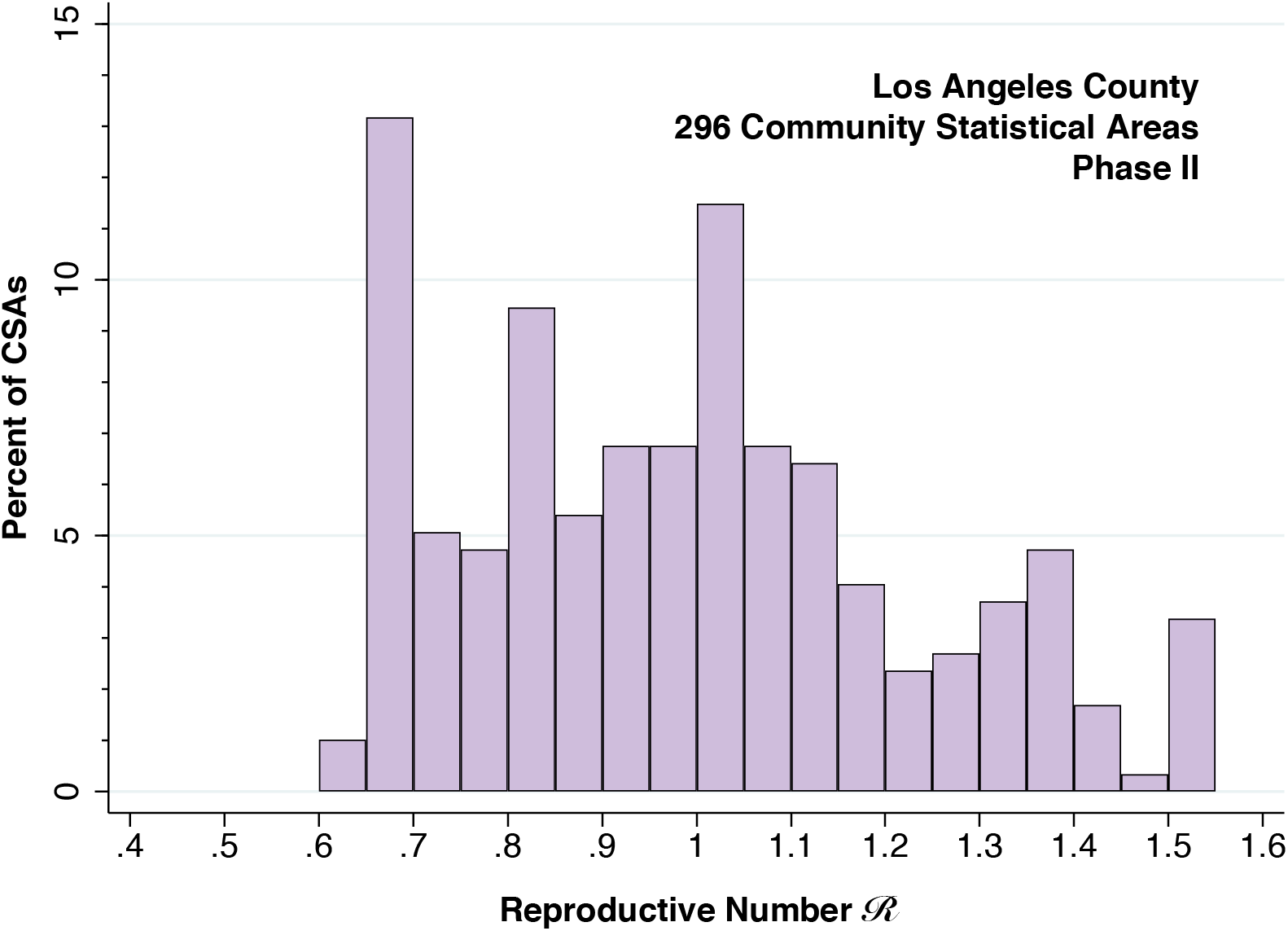
Distribution of Reproductive Number ℛ Among 296 Countywide Statistical Areas During Phase II

While the lockdowns reduced ℛ in the aggregate from about 2.7 in Phase I to 1.0 in Phase II, Figure 5 informs us that the velocity of the epidemic in Phase II remained quite variable. As we noted in connection with our discussion of Figure 1, county-level officials reopened retail stores, indoor dining, hair salons, gyms and bars in May and June during Phase II. Figure 5 suggests that the impact of such reopening was likely to be quite variable.

### 3.4. Phase III: The Paradox of the Negative Coefficient

Table 5 displays the spatial model results for Phase III. The reduced estimates of the transmission parameter *α* in Models 0 and 1 are consistent with an inhibitory effect of the California health officer’s order reversing most of the county’s prior reopening orders of Phase II (Gutierrez 2020). The results of Model 2, however, tell a different story. The spatial influence parameter *γ* is now insignificant, while the coefficient *δ* of *MULTI* is now significantly *negative*. The right-hand panel of Table 5, displaying the alternative case where *f* = 0.5, shows that this finding is not an artifact of our assumption concerning the extent of underascertainment of confirmed cases.

The apparent paradox of the negative coefficient is resolved in Figure 6, which shows the time paths of estimated proportions of infective *I*_*it*_ and susceptible *S*_*it*_ in two groups of CSAs: those in the lower half and those in the upper half of the distribution of *MULTI*. The estimates are derived from the alternative case where *f* = 0.5. Those CSAs with a higher proportion of at-risk multi-generational households, rendered as the darker blue curves, consistently had a higher prevalence of active infection. However, after the state’s reversal order at the end of Phase II, the proportion infected declined *more rapidly* among the high-*MULTI* CSAs. The inference is that the renewed lockdown had a greater deterrent impact on social mobility in those communities with a higher proportion of multi-generational households.

**Figure 6.**
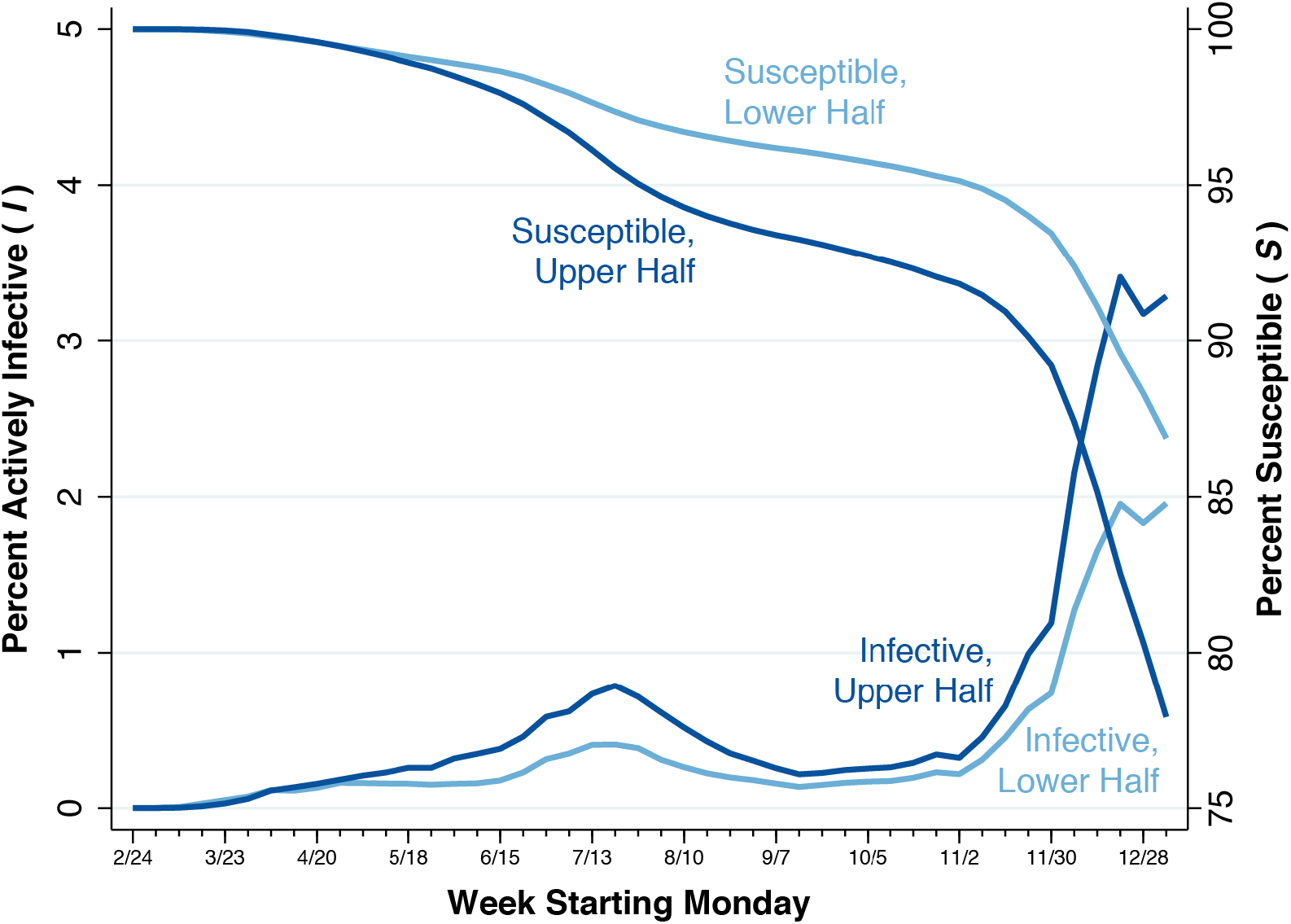
Time Path Percent Infective (I) and Percent Susceptible (S) in CSAs the Upper and Lower Halves of the Distribution of MULTI

Figure 7 below relies upon smartphone mobility data to further explore the basis for this conclusion. Here, we’ve broken down CSAs into four quartiles of the distribution of *MULTI*, with the highest quartile rendered as the darkest curve. For each of the four groups of CSAs, the curve shows the trend in the number of fast-food-restaurant visits by smartphones originating in CSAs within that group. The trends have been normalized so that the mean number of visits during the weeks of February 10 and 17, before the start of our study period, are equal to 100. Truncated at the left end of the graph are the precipitous declines in restaurant visits in all four groups during Phase I. Thus, the time line starts with the week of March 30 (week 5).

**Figure 7.**
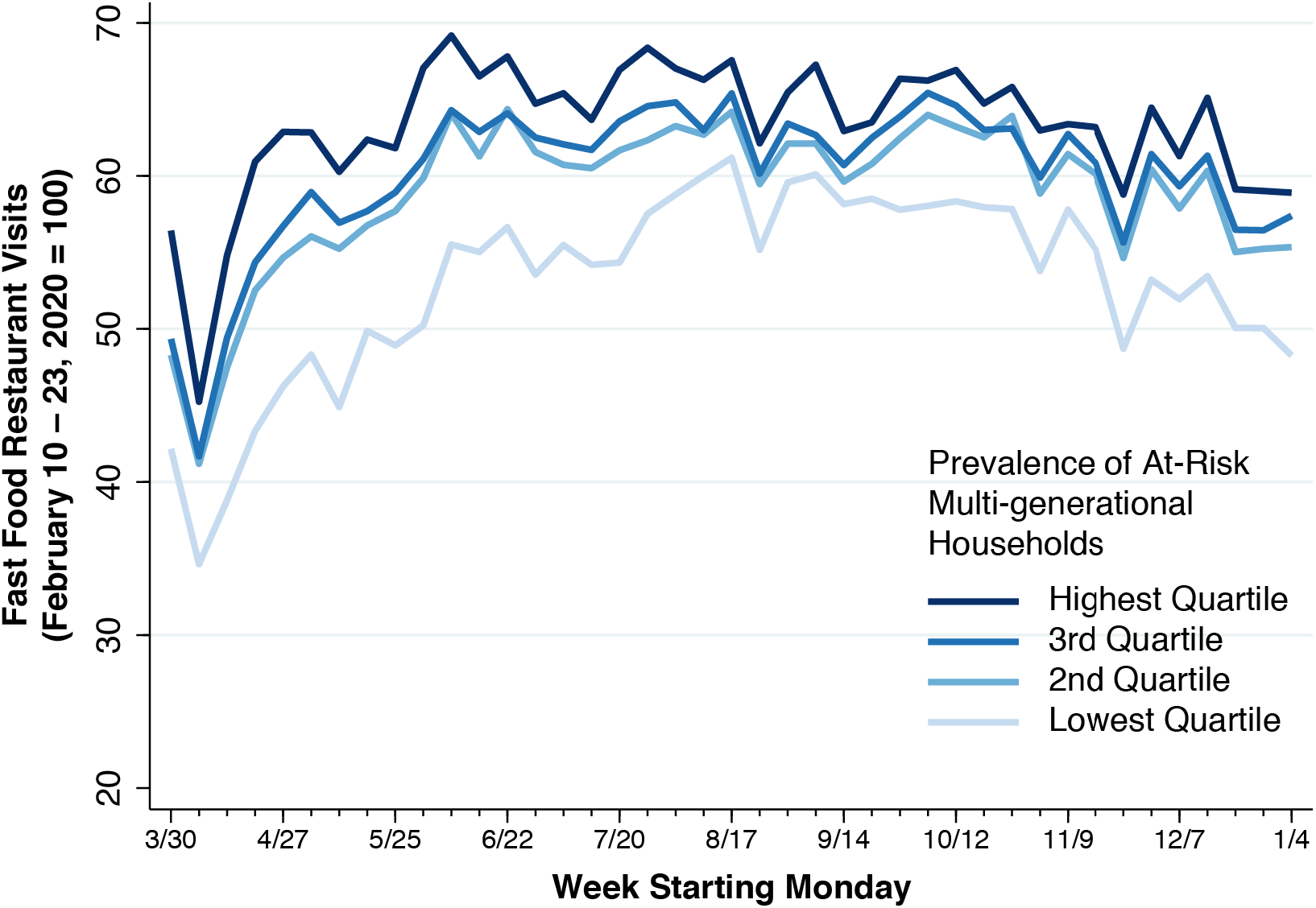
Visits to Fast Food Restaurants from Smartphones Originating in CSAs in the Four Quartiles of the Distribution of MULTI

Figure 7 shows all four quartiles of *MULTI* reaching the nadir of fast-food-restaurant visitation rates during the week of April 6 (week 6). By that point, CSAs in the lowest quartile had exhibited the largest declines in response to the lockdowns of Phase I. Visitation rates recovered in all four groups during Phase II, as local restrictions were relaxed. Once the state health officer reversed these local reopening orders during the week of July 13 (week 20), visitation rates in the higher quartiles began to turn around, but the lowest quartile did not. By the week of September 14 (week 29 in Phase III), the spread between the highest and lowest quartiles had narrowed to less than 5 percentage points.

### 3.5. Phase IV: The Reproductive Number ℛ Climbs Back Up To 1.4

Table 5 below shows our spatial model estimates for Phase IV. The most salient feature of Phase IV is the marked rise in the reproductive number ℛ back up to 1.4, despite the emergency stay-at-home orders issued by the Los Angeles mayor and the California regional health officer during week 40 (Garcetti 2020b, Pan 2020). As in Table 4, we show the alternative case where the underascertainment proportion *f* is assumed to be 50 percent. The insignificant coefficient of the radial expansion parameter *γ*, also seen in Model 2 for Phase III, indicates that within-CSA transmission had become the dominant mode of viral propagation.

While the estimated coefficient *δ* of *MULTI* is smaller than the estimates for Phases I and II, it remains significantly different from zero. Thus, a 12-percentage-point increase in the prevalence of at-risk multi-generational households, equivalent to the difference between the population-weighted mean values of *MULTI* in the top and bottom half of the distribution, would increase the transmission parameter by only about 12 *γ* ≈ 0.11. Still, over the course of the 11 weeks of Phase IV, that increment alone would be sufficient to account for the divergence in the prevalence of active infection seen in Figure 6.

To bring home this point, Figure 8 plots the relation between the estimated cumulative proportion of infections (in the notation of our spatial model, 1 – *S*_*it*_ at *t* = 45) against the prevalence of at-risk multi-generational households (that is, *Z*_*i*_). As in Figure 6, the estimates are derived from the alternative case where *f* = 0.5. The fitted line has a slope of 0.777 (95% confidence interval, 0.678–0.875). With the exception of some outlier CSAs with relatively small populations, the communities with the highest values of *MULTI* had a predicted cumulative prevalence approaching one-third of the population. Communities with the lowest values of MULTI, by contrast, had a predicted cumulative prevalence under 10 percent. The predictions of the spatial SIR model in Figure 8 are thus broadly consistent with the dispersion in the cumulative incidence of confirmed cases seen in Figure 2.

**Figure 8.**
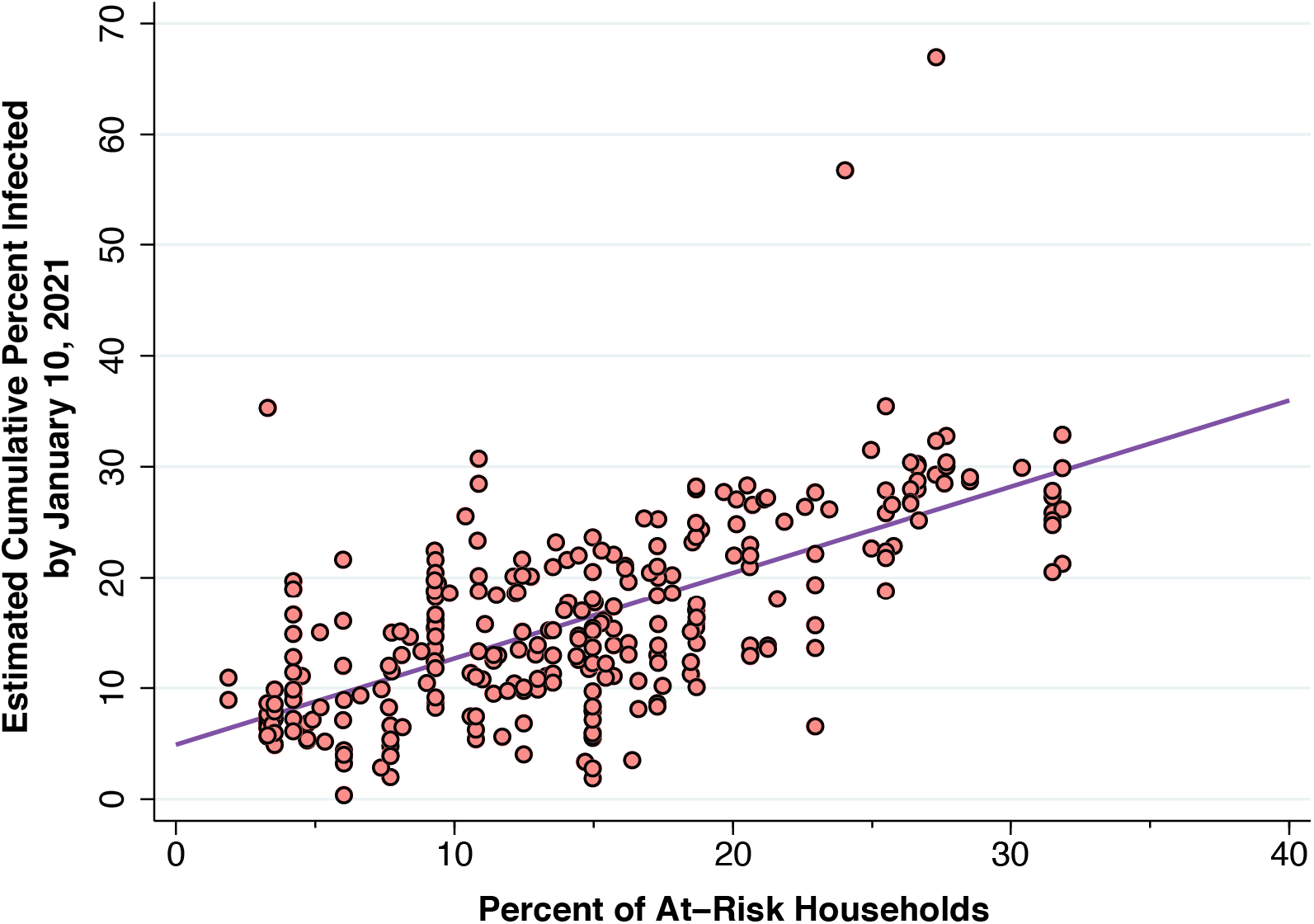
Predicted Cumulative Incidence of Infection (1 – S) Through Week 45 in Relation to the Proportion of At-Risk Multi-Generation Households (MULTI)

While Figure 8 helps to explain the striking geographic variation in the burden of new SARS-CoV-2 infections during Phase IV, it does not address the underlying causes for the marked increase in the overall reproductive number to 1.4. If the increase in ℛ were attributable to lapses in compliance with social distancing policies, we would have expected to see an increase in restaurant visits during the final weeks in Figure 7 above. Neither has an increase in the amount of time spent outside the home been observed (Harris 2020f).

## Discussion

### Summary of Findings

We identified four phases of the epidemic in Los Angeles County during February 24, 2020 through January 10, 2021 (Figure 1). Phase IV (running for 11 weeks starting October 19) accounted for more than two-thirds of all confirmed COVID-19 cases during the entire 46-week interval under study.

The map of the cumulative incidence of confirmed infections during Phase IV, we found, bore a striking resemblance with the corresponding map of the prevalence of at-risk multi-generational households (Figure 2). In a multivariate cross-sectional analysis of approximately 300 communities in the county, the prevalence of at-risk multi-generational households (*MULTI*) was a more important determinant of the cumulative incidence of confirmed infections in Phase IV than other community-specific indicators. These included the proportion of households with a total income below that of a single minimum-wage worker (*INC22*), the proportion of households with at least one worker in a low-wage occupation that could not be performed at home (*OCCUP*), and the proportion of households dependent on food stamps (*SNAP*). (Figure 3 and Table 2).

We formulated a spatial modification of an SIR epidemic model that tested two distinct multiplicative effects on the transmission rate in each community: the effect of infection rates in adjacent communities (the parameter *γ*), and the effect of multi-generational household prevalence within the same community (the parameter *δ*). We estimated this model separately on the data for each of the four phases.

Phase I (weeks 0–5) was characterized by substantial adjacent-community effects operating within a narrow radius of 1 kilometer (Table 3). This finding was in accordance with serial weekly maps of the spread of infection during Phase I, which showed initial, rapid radial extension from a focus originating in an affluent area containing such communities as Beverly Crest (Figure 4). Phase I was also characterized by the significant influence of within-community multi-generational prevalence on transmission rates. As the epidemic expanded, hotspots began to develop in areas with higher concentrations of multi-generational households. The estimated overall reproductive number ℛ during this phase was on the order of 2.7 (Table 3).

The flattening of the epidemic curve in Phase II (weeks 6–20) reflected the effects of voluntary and coerced social distancing, particularly the state-of-emergency orders issued during Phase I. Still, both adjacent-community effects and the within-community impact of high proportions of multi-generational households remained significant (Table 4). While the global, countywide reproductive number ℛ hovered around 1.0, local reproductive numbers varied widely from 0.6 to 1.5, depending on the prevalence of multi-generational households in the community (Figure 5).

As the epidemic curve began to rise toward the end of Phase II, the state ordered the reversal of measures taken at the county level to easy restrictions on retail stores, indoor dining, hair salons, gyms and bars (Figure 1). As confirmed case incidence declined during Phase III (weeks 21–34) in response to this public policy intervention (Figure 1), the countywide reproductive number ℛ dropped to about 0.6 and adjacent-community effects were no longer consistently detectable (Table 5). More striking, however, was the finding that a high prevalence of multi-generational households was significantly associated with decreased – rather than increased – viral transmission (Table 5). Phase III, it turned out, saw a narrowing in the gap in infection rates between communities with high and low percentages of multi-generational households (Figure 6). This interpretation was supported by smartphone tracking data showing that the gap in restaurant visitation rates during this period had also narrowed (Figure 7).

During Phase IV (weeks 35–45), infect rates surged, with the global reproductive number ℛ reverting to 1.4 (Table 5). Spillover effects between communities were no longer detectable. The model parameter (*δ*) relating multi-generational household prevalence to the transmission rate, while smaller than at the start of the epidemic, was still sufficiently large to generate a wide dispersion in cumulative infection rates. By the end of our study period (the week of January 4, 2021), estimated cumulative infection rates varied from under 10 percent in communities with low multi-generational prevalence to greater than 30 percent in communities with a high percentage of at-risk multi-generational households (Figure 8).

These findings, taken together, supported a critical role of household structure in the initial dissemination and continued wide propagation of SARS-CoV-2 infection in Los Angeles County.

### Strengths and Limitations of This Study

Our study takes advantage of the cohort structure of our database, in which we follow a group of related geographic units longitudinally over time. This structure allowed us to test a model of radial geographic expansion during Phases I and II of the Los Angeles County COVID-19 epidemic, and to elucidate the substantial heterogeneity of transmission patterns within the county over time. While the global reproductive number ℛ hovered around 1 during Phase II, our approach permitted us to discern local reproductive numbers ranging from 0.6 to 1.5. While the overall confirmed case incidence rate rose by about 10-fold during Phase IV, we were able to identify wide community-specific dispersion in cumulative disease rates.

On the other hand, our study is exclusively population-based. We do not follow a longitudinal cohort of individual households to see how many young adult members went to a restaurant or a gym, got infected, and then brought their infections home to older household members. A population-based indicator such as the proportion of households at risk for multigenerational transmission (*MULTI*) could thus be criticized as no more than a proxy for some other correlated characteristic of the community.

To be sure, we confirmed the quantitative importance of multi-generational household prevalence in a cross-sectional regression analysis that included other measures of poverty (*INC22, SNAP*) as well as the proportion of households with high-risk workers (*OCCUP*) (Table 2). Still, one might posit that the critical underlying variable is the proportion of Spanish-speaking households with uninsured members (Weng, Saal, and Chan 2020, Vijayan et al. 2020). One might similarly contend that our variable *MULTI*, which relied on the presence of at least one younger adult (aged 18–34) and another older adult (aged 45 or more) in the household, was no better an indicator of multi-generational transmission risk than, say, the number of persons per bathroom in the household. Data on the prevalence of such risk factors as smoking, elevated body mass index and comorbidities such as diabetes have been considered (Horn et al. 2020), but these cofactors are more relevant to a study of disease morbidity and mortality.

Our principal endpoint was the incidence of confirmed cases of COVID-19. It is now widely acknowledged that confirmed case counts significantly understate the actual numbers of SARS-CoV-2 infections (Havers et al. 2020).While we adjusted our spatial SIR model to account for an estimated 40–50 percent underascertainment, there is some evidence that underascertainment rates are much higher (Wu et al. 2020, Sood et al. 2020). While we estimated that cumulative infection rates ranged from 10 percent to upwards of 30 percent across communities, another unpublished model suggested that one in three residents of Los Angeles had already been infected (County DHS COVID-19 Predictive Modeling Team 2021).

One alternative endpoint would be seroprevalence, but serial population-based studies of seroprevalence are still uncommon (Hallal et al. 2020), and there is evidence that population seroprevalence may decline with time (Buss et al. 2020). Hospital admission rates have been studied as an alternative to confirmed case incidence (Harris 2020b, g), but such an endpoint would also depend on case severity. The test positivity rate – the proportion of positive tests among all persons tested – has been employed as an endpoint in cross-sectional studies (Vijayan et al. 2020, Cotti et al. 2020). Adaptation of this endpoint to a dynamic epidemic model is problematic, however, as the number of individuals tested is endogenous and must be modeled as well (Bhaduri et al. 2020).

To explain the paradoxically negative value of the parameter *δ* during Phase III, we relied on data on smartphone visits to fast-food restaurants as an indicator of social mobility (Figure 7). While data on smartphone visits to restaurants and bars have been repeatedly used as a measures of potential coronavirus exposure (Harris 2020b, e), there has been no independent verification of their accuracy.

Our specification of a spatial modification of the conventional SIR model adheres to the modeling philosophy that one should introduce the minimum necessary modifications of the most parsimonious model (Harris 2020b). One might contend that the appropriate base model would instead be the SEIR (susceptible-exposed-infective-resistant) version, which has been widely employed in studies of SARS-CoV-2 transmission (Radulescu, Williams, and Cavanagh 2020, Li et al. 2021, Godio, Pace, and Vergnano 2020). It is hardly clear, however, that the problem of coming up with the additional between-state transition parameters in the SEIR model is any more tractable than our problem of devising an inflation factor to account for unascertained cases in Section 2.5 above.

### Characterizing the Initial Outbreak

Our geospatial mapping study (Figure 4), in combination with our estimates of a spatial SIR model for Phase I (Table 3), permitted us to characterize the initial outbreak of COVID-19 in Los Angeles County. The earliest days of the outbreak saw multiple parallel importations in several relatively affluent areas of the county where residents had the resources to travel. A phylogenetic analysis of SARS-CoV-2 samples drawn during March 22 – April 15 at a major hospital located within one of the initial foci of infection found that the larger proportion belonged clades derived from Europe (Zhang et al. 2020). The epidemic then spread by radial expansion over a period of weeks from this focus of infection to nearby communities with a higher prevalence of multi-generational households. This pattern of spread by radial extension stands in sharp contrast to the earliest days of the outbreak in New York City, where community-transmitted infections were dispersed throughout all five boroughs in a matter of days (Harris 2020c, Gonzalez-Reiche et al. 2020). Our estimate of a reproductive number ℛ equal to 2.7 (Table 2, base case, Model 2) is consistent with the estimate of ℛ_0_ in the range of 2.43–3.10 for the Italy (D’Arienzo and Coniglio 2020), but falls below the estimates of 3.47 (range, 3.16–3.78) for New York City (Harris 2020d) and 3.54 (range, 3.40–3.67) for Wuhan (Hao et al. 2020), both of which had massive subway systems.

### Heterogeneous Responses to Public Policies

Our analysis of a longitudinal panel of diverse communities within Los Angeles County helps us understand how responses to epidemic-control policies can be so heterogeneous. After the state-of-emergency orders issued in March, the county entered into Phase II during the week of April 6 with a global reproductive number ℛ of 1.0. Yet Figure 5 informs us that the local reproductive numbers varied widely from 0.6 to 1.5.

In response to the flattening of the global epidemic curve in Phase II, county policy makers reopened retail stores, indoor dining, hair salons, gyms and bars in May and June (Parvini 2020, Shalby 2020a, Money 2020). When these actions overshot the mark, the Los Angeles County health officer issued an order closing indoor onsite dining (Los Angeles County Department of Public Health 2020). On July 13, the state public health officer closed indoor operations in bars not concurrently serving meals, as well as gyms in counties on its monitoring list, to which Los Angeles County already belonged (Angell 2020).

All of these public policy decisions applied to the entirety of Los Angeles County. Yet the evidence is that the responses to these policies varied widely. Figure 7 shows that fast-food restaurant visits by residents of those communities in the top quartile of multi-generational household prevalence, which had the highest reproductive numbers, had a very different response than visits by residents of communities in the lowest quartile, which had the lowest reproductive numbers. While this finding alone does not establish that all indicators of social mobility responded in the same manner, it highlights the bluntness of policy instruments that were to be applied uniformly to a county of 10 million inhabitants.

### What Caused the Phase IV Surge?

While Figures 3 and 8 and Table 2 establish the substantial contribution of multi-generational households to the surge in confirmed case incidence observed in Phase IV, they do not tell us why the surge occurred in the first place. In terms of our spatial SIR model, they do not explain the striking rebound in the transmission parameter *α* that is evident in the data.

Figure 7 tells us that visits to fast-food restaurant visits declined overall during Phase IV. Other smartphone-based indicators of social mobility showed little change (Harris 2020f) despite the emergency stay-at-home orders issued by the Los Angeles mayor and the California regional health officer during week 40 (Garcetti 2020b, Pan 2020). Even visits to gyms, compiled in an earlier draft of this article (Harris 2020h), continued to decline.

Two plausible explanations come to mind. First, smartphone-based indices of social mobility have not captured large family gatherings that occurred during the succession of winter holidays that began with Thanksgiving. The publicly available smartphone data show only how frequently individual device holders moved and where they went, but not how many were congregated in the same place. A subsequent decline in the frequency of such high-density gatherings may help to explain the drop in confirmed case incidence that has so far been observed in January 2021, outside the observation interval of the present study. Second, a new strain of SARS-CoV-2 could have emerged in Southern California (Zhang et al. 2021). The difficulty with the latter explanation is that it does not readily explain the subsequent post-Phase IV decline in incidence that appears now to be under way.

### Implications

Despite an array of aggressive public policies aimed at reducing social mobility, our findings suggest that intra-household transmission has been a critical vehicle for the persistence of the COVID-19 epidemic in Los Angeles County. The prevalence of at-risk households in a community, it appears, is not simply a predictor of the persistence of coronavirus transmission, but also a multiplier of the effects of other policies aimed at social distancing. The impact of preventing one case of asymptomatic infection in a socially active young adult, who would otherwise have brought his or her infection into the household, will depend directly on the number of susceptible household members who have been spared.

Our results cast a pessimistic shadow on so-called targeted policies that selectively relax restrictions on lower-risk, younger persons while seeking to protect more vulnerable older persons (Chikina and Pegden 2020, Acemoglu et al. 2020, Iverson, Karp, and Peri 2020, Gollier 2020). Such a policy might be feasible in settings where older persons are sequestered in retirement communities or assisted living facilities, but the data here show that this is not the reality of Los Angeles County.

Most importantly, our findings require us to view the household rather than the individual as the foremost target of healthcare policy. The message “protect yourself” (*protégete* in Spanish) needs to be reconfigured as “protect your family” (*protege a tu familia*). When a healthcare provider encounters a new patient with suspected or established COVID-19, the interview needs to turn quickly to questions about other household members, their health status, and their symptoms. The widely recognized model of the patient-centered medical home (Alexander and Bae 2012) needs to be replaced by the family- and household-centered medical home.

## Data Availability

The author will make available all data, programs, and output.

## Footnotes

* For an earlier version of this study (Harris 2020h), we relied on DPH press releases to reconstruct community-specific case counts. The present study relies upon a more extensive, updated database issued by the DPH.

† In the earlier version of this study (Harris 2020h), we relied on the 2018 installment of the ACS. The present study relies upon the more recently released 5-year 2015–2019 installment.

‡ The occupation codes included: 3601 Home Health Aides; 4020 Cooks; 4030 Food Preparation Workers; 4055 Fast Food And Counter Workers; 4120 Food Servers, Nonrestaurant; 4140 Dishwashers; 4220 Janitors And Building Cleaners; 4230 Maids And Housekeeping Cleaners; 4251 Landscaping And Groundskeeping Workers; 4255 Other Grounds Maintenance Workers; 6260 Construction Laborers; 6600 Helpers, Construction Trades; 7840 Food Batchmakers; 9350 Parking Lot Attendants; 9640 Packers And Packagers, Hand; 9645 Stockers And Order Fillers; and 9720 Refuse And Recyclable Material Collectors.

§ These chains included: Arby’s, Carl’s Jr., Chick-fil-A, Five Guys, Jack in the Box, Johnny Rockets, Jollibee, McDonald’s, Panda Express, Rally’s, Subway, Wendy’s, and Wienerschnitzel.

